# Visual Memory Performance Correlates with Alzheimer’s Disease Biomarkers in Cognitively Normal Individuals

**DOI:** 10.1101/2025.06.27.25330448

**Authors:** Taylor A. James, Liping Zhao, Han Xu, Junjie Wu, David A. Schweidel, Felicia C. Goldstein, Allan I. Levey, Deqiang Qiu, David W. Loring, John J. Hanfelt, James J. Lah

## Abstract

**INTRODUCTION:** Alzheimer’s disease (AD) pathology evolves silently for many years prior to disease, and early identification of individuals at greatest risk is critical for improving outcomes.

**METHODS:** We conducted exploratory factor analysis (EFA) using neuropsychological testing from 1,697 healthy adults. Factor scores were submitted as predictors for structural MRI, functional connectivity, and cerebrospinal fluid (CSF) biomarkers.

**RESULTS:** The EFA model identified five latent factors. Factor 1, Verbal Memory, included high loadings from Rey Auditory Verbal Learning Test, and Factor 2, Visual Memory, included high loadings from Rey Complex Figure Test (RCFT). Factor 2 positively correlated with hippocampal volume, precuneus and posterior salience network connectivity, and negatively with CSF tau. Factor 1 correlated with hippocampal volume but not with network connectivity or CSF biomarkers.

**DISCUSSION:** Results demonstrate a significant relationship of visual memory with AD biomarkers. RCFT may be an important assessment for evaluating and tracking individuals during the earliest stages of Alzheimer’s pathology.

## 1. Introduction

Alzheimer’s disease (AD) is a progressive neurodegenerative disorder, characterized by gradual cognitive decline, leading to loss of independence and ultimately death. The disease progresses along a continuum, beginning with an asymptomatic phase lasting up to two decades, during which pathophysiological changes are detectable but cognitive symptoms are not. Eventually, these changes can give rise to the symptomatic phases manifesting as mild cognitive impairment (MCI) and progressing to increasingly severe stages of dementia. The transition from asymptomatic to symptomatic stages represents a critical period for intervention; by the time symptoms are clinically observable, significant neuronal damage has already occurred.^1^ While current treatments show promise for slowing disease progression in individuals at early stages of disease,^2,3^ they are ineffective if administered after significant neurodegeneration has taken place.^4^ Recognizing the need for earlier interventions, research has increasingly focused on identifying biomarkers and imaging techniques capable of detecting AD pathology in the asymptomatic phase, allowing for interventions that could delay or prevent clinical disease.

The progression of AD pathology has been well-documented, with amyloid-beta (Aβ) plaques and neurofibrillary tau tangles being identified as core hallmarks of the disease. Aβ deposition precedes neocortical tau pathology, with amyloid accumulation beginning many years before the onset of cognitive impairment.^5^ In the asymptomatic phase, elevated amyloid burden is detectable through positron emission tomography (PET) imaging, cerebrospinal fluid (CSF), or blood-based biomarkers, despite an absence of clinical symptoms.^6^ Following amyloid deposition, hyperphosphorylated tau proteins begin to form tangles, leading to neuronal damage, brain atrophy, and eventually cognitive impairment.^7,8^ The presence of these pathological changes in asymptomatic individuals suggests a lengthy window during which early intervention could, in theory, delay or prevent progression to symptomatic AD.^1^

Amyloid PET imaging and CSF assays for Aβ and tau are the two best established tools for identifying amyloid pathology in non-demented individuals.^9^ Additionally, presence of the *APOE ε4* allele can identify those at heightened genetic risk.^10^ Neuroimaging methods assessing brain structure and connectivity, such as structural MRI and functional MRI (fMRI), have also been explored as predictors of AD progression. Structural MRI can detect early atrophy in regions like the hippocampus and entorhinal cortex, key sites of early neurodegeneration in AD,^11-13^ while fMRI can reveal disruptions in brain network connectivity that precede cognitive impairment.^6,14^

In parallel with the development of biomarker-based frameworks, considerable effort has been directed toward identifying sensitive cognitive measures that can detect early signs of AD-related decline in asymptomatic individuals. One prominent example is the Preclinical Alzheimer Cognitive Composite (PACC)^15^, which combines episodic memory, executive function, and global cognition to track subtle cognitive decline prior to symptom onset. The PACC has since been widely adopted in preclinical AD trials and shown to be sensitive to amyloid-related changes in cognitively normal individuals.^16,17^ In addition to composite measures, longitudinal studies have explored the absence of learning curves or diminished practice effects as potential early indicators of progression to MCI or AD.^18,19^ These efforts underscore the potential value of neuropsychological testing for detecting incipient AD-related decline.

The goal of the current study is to determine whether neuropsychological performance in different cognitive domains among cognitively unimpaired individuals can predict early markers of AD (i.e., reduced brain volume in key regions, disruptions in functional connectivity, and CSF levels of Aβ and tau). To accomplish this goal, we conducted an exploratory factor analysis (EFA) on baseline neuropsychological testing data collected on 1,697 Emory Healthy Brain Study (EHBS) participants.^20^ The factor analysis allowed for the grouping of tests into cognitive domains. The factors representing these cognitive domains were then used as predictors in median regression models to evaluate their relationship with structural MRI, resting state functional connectivity, and CSF Aβ and tau.

## 2. Method

### 2.1. Participants

For a participant to qualify for EHBS,^20^ they must be between the ages of 50 and 75 with no preexisiting diagnosis of MCI, dementia, or other cognitive impairment, be willing to travel to Emory, and be able and willing to complete a lumbar puncture, brain MRI, and other study procedures. Exclusionary criteria for enrollment include being unable to speak or understand English, current use of anticoagulant medication, history of bleeding disorders, stroke or transient ischemic attack, congestive heart failure, multiple sclerosis, amyotrophic lateral sclerosis, Parkinson’s disease, epilepsy, active HIV, tuberculosis, hepatitis B or C virus not fully treated, schizophrenia, kidney disease requiring dialysis, solid organ transplant, and life-threatening illnesses. The protocol was approved by the Emory Institutional Review Board and all participants provided written informed consent. The study was performed in accordance with the ethical standards in the 1964 Declaration of Helsinki.

Data used in the current analyses are from 1,697 EHBS participants’ baseline visits, completed prior to March 17, 2025. Mean age of participants at their baseline visit was 63.0±6.8 years and mean education was 16.6±2.1 years, indicating most participants had at least a four-year degree. Other demographic characteristics (sex, race, and ethnicity) are presented in **Table 1** along with *APOE* genotypes.

**Table 1.**
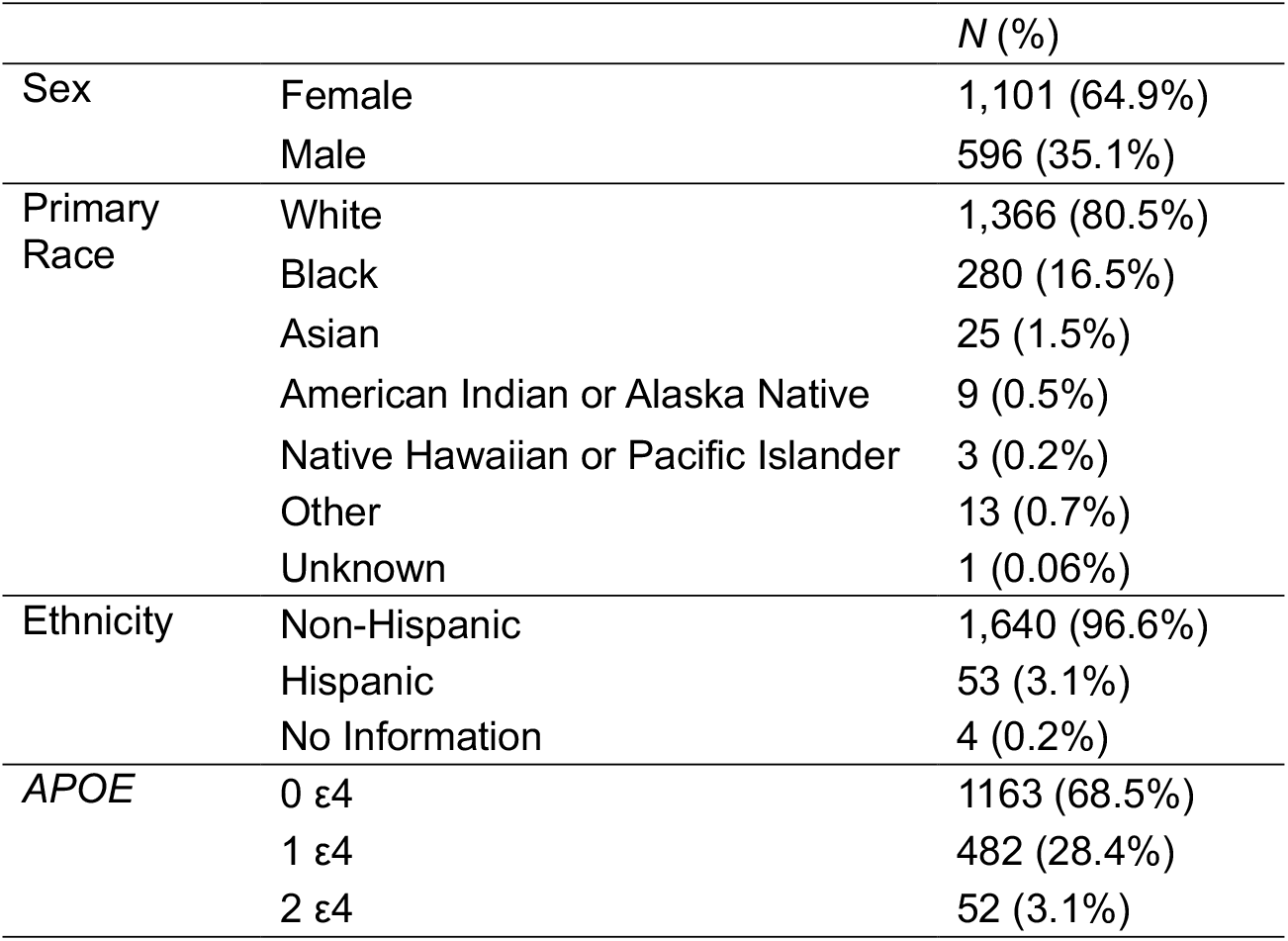
Participant Demographics and APOE Genotypes.

### 2.2. Procedures

Participants provided details on their medical history including current medications and completed several surveys including the Patient Health Questionnaire (PHQ-8),^21^ Generalized Anxiety Disorder scale (GAD-7),^22^ Cognitive Function Instrument (CFI),^23^ and Quick Dementia Rating System (QDRS).^24^ The EHBS neuropsychological battery consisted of the Montreal Cognitive Assessment (MoCA),^25^ Rey Auditory Verbal Learning Test (RAVLT),^26^ Rey Complex Figure Test (RCFT),^27,28^ Number Span Test Forward (NSF) and Backward (NSB),^29^ Judgement of Line Orientation (JOLO),^30^ Controlled Oral Word Association Test (COWA),^31^ Category Fluency for animals,^32^ Multilingual Naming Test (MiNT),^33^ Trail Making Test (TMT) A and B,^34^ and the NIH Toolbox Picture Vocabulary Test.^35^

Several changes in the neuropsychological battery impacted data availability. From April 2017 to August 2018, the Free and Cued Selective Reminding Test^36,37^ was used in place of RAVLT. For COWA, F fluency was always collected as part of the MoCA; FAS fluency was used until March 2021, when it was replaced by FL fluency. TMT and the NIH Toolbox Picture Vocabulary Test were administered only during in-person visits, and restrictions on in-person activities during the COVID-19 pandemic limited data collection.

MoCA was not used in the factor analysis, as it measures a variety of cognitive processes. NIH Toolbox Picture Vocabulary Test was not used, as 23% of participants were missing data, and MiNT was not used, as performance neared ceiling for most participants (mean = 30.6±1.9 out of 32 total points).

### 2.3. Analyses

#### 2.3.1. Statistical Analysis

We analyzed the EHBS baseline data to explore the relationship of neuropsychological performance with MRI regional brain volumes, resting-state functional connectivity, and with CSF biomarkers using R software (v4.3.1). Correlations of cognitive domain factor scores with outcome variables were evaluated with median regression models using the *quantreg* package (v5.96),^38^ while controlling for demographic variables (age, education, sex, and race) and presence or absence of an *APOE ε4* allele. To correct for multiple comparisons, Benjamini-Hochberg (B-H) procedure was applied, and a B-H critical value for a false discovery rate of 0.20 was chosen. The p values were sorted and compared with their corresponding B-H critical values for each factor score.

#### 2.3.2. Missing Data Mechanism

Because data were missing on some neuropsychological assessments, the missing data mechanism was diagnosed. There was no systematically missing data based on the data collection procedure and clinical judgement. The nonparametric test of homoscedasticity across different patterns of missing data in the *MissMech* package (v1.0.3)^39^ was used to check if data were missing completely at random (MCAR). The hypothesis of MCAR was not rejected at 0.05 significance level (*p* = .443). We also checked whether missing data were related to demographic and cognitive variables. Compared to the group with no missing data, those missing data had lower education (16.2 ± 2.2 vs. 16.8 ± 2.0, *p* < .001). No other demographic variables (*p*s > .08) nor any cognitive variables (*p*s > .06) significantly differed between the groups.

To increase statistical power and obtain robust estimates accounting for uncertainty introduced by missing data, we performed multiple imputation to predict missing values in each incomplete cognitive score based on observed data and demographic variables. The multiple imputation was conducted with Multivariate Imputation with Chained Equations (MICE) from the *mice* package (v3.16.0),^40^ then the covariance matrices of the imputed data sets were combined into a single covariance matrix using Rubin’s rules in the *mifa* package (v0.2.0).^41^ Nonparametric bootstrap confidence intervals were generated for the variance explained by different numbers of principal components. Thirty imputed datasets were created with age, gender, race, and education as predictors in the imputation models. The resulting covariance matrix was converted to a correlation matrix for the EFA.

#### 2.3.3. Exploratory Factor Analysis

We conducted EFA using the *EFATools* package (v0.4.4).^42^ First, we used the Kaiser-Meyer-Olkin (KMO) test as a measure of sampling adequacy and Bartlett’s test of sphericity to determine the factorability of the data. Next, the determinant score was calculated to examine multicollinearity among the variables. To determine the number of factors to retain, we employed three methods: the Kaiser-Guttman rule (or Kaiser’s criterion; retain all factors with associated eigenvalues > 1), examination of the scree plot, and parallel analysis using EFA and means decision rule. Finally, we performed EFA using principal axis factoring with a promax rotation to facilitate interpretation of results. An oblique rotation was selected, as factors were expected to correlate with one another. Variables were assigned to factors if their loadings exceeded 0.30 and loaded on a single factor.

Bartlett factor scores were created to evaluate the relationships between the factors and MRI volumetric data, resting state functional connectivity, and CSF biomarkers (amyloid-beta 42 [Aβ42], phosphorylated tau-181 [pTau], and total tau [tTau]) in median regression models. CSF samples were assayed on the Roche Elecsys platform^43^ with standard pre-processing procedures as described.^44^ Details on MRI acquisition, data preprocessing, and identification of functional networks are reported elsewhere (Wu *et al*., submitted). We used 14 functional networks from templates identified by Shirer *et al*.^45^: auditory, basal ganglia, left (LECN) and right (RECN) executive control, language, precuneus, sensorimotor, visuospatial, anterior salience, posterior salience, dorsal and ventral default mode (DMN), higher visual, and primary visual. For the structural MRI data, we analyzed bilateral AAL regions for the hippocampus and entorhinal cortex. The structural MRI data were corrected for the total intracranial volume (proportion of total brain volume).

## 3. Results

### 3.1. Exploratory Factor Analysis

Correlations between variables used in the factor analysis are presented in **Table 2**. While some strong correlations exist between variables (e.g., RAVLT immediate and delayed recall, *r* = .84, RCFT immediate and delayed recall *r* = .91), the determinant of the correlation matrix was 0.0006, suggesting multicollinearity was not a significant concern for these data. The overall KMO value was 0.831, and the Bartlett’s test of sphericity was significant, χ^2^(120) = 12,535.09, *p* < .001, suggesting the data are suitable for factor analysis.

**Table 2.** Correlations between Neuropsychological Test Scores.

|  | RAVLT<br>A1-5 | RAVLT<br>A6 | RAVLT<br>A7 | RAVLT<br>Rec | NSF | NSB | JOLO | F<br>Flu | L<br>Flu | Cat<br>Flu | RCFT<br>Copy | RCFT<br>IR | RCFT<br>DR | RCFT<br>Rec | TMT<br>A | TMT<br>B |
| --- | --- | --- | --- | --- | --- | --- | --- | --- | --- | --- | --- | --- | --- | --- | --- | --- |
| RAVLT A1-5 | 1 | .79 | .78 | .58 | .18 | .29 | .15 | .23 | .27 | .34 | .24 | .28 | .29 | .18 | -.23 | -.29 |
| RAVLT A6 |  | 1 | .84 | .61 | .12 | .24 | .14 | .18 | .21 | .33 | .21 | .30 | .30 | .16 | -.22 | -.26 |
| RAVLT A7 |  |  | 1 | .64 | .09 | .20 | .14 | .21 | .23 | .31 | .21 | .30 | .29 | .16 | -.22 | -.24 |
| RAVLT Rec |  |  |  | 1 | .04 | .16 | .09 | .14 | .17 | .22 | .18 | .23 | .24 | .14 | -.17 | -.18 |
| NSF |  |  |  |  | 1 | .45 | .16 | .17 | .25 | .13 | .08 | .11 | .12 | .06 | -.16 | -.21 |
| NSB |  |  |  |  |  | 1 | .23 | .19 | .23 | .23 | .17 | .20 | .19 | .12 | -.17 | -.27 |
| JOLO |  |  |  |  |  |  | 1 | .10 | .18 | .23 | .35 | .32 | .31 | .19 | -.27 | -.33 |
| F Flu |  |  |  |  |  |  |  | 1 | .57 | .35 | .10 | .12 | .13 | .07 | -.20 | -.25 |
| L Flu |  |  |  |  |  |  |  |  | 1 | .39 | .18 | .16 | .16 | .09 | -.24 | -.32 |
| Cat Flu |  |  |  |  |  |  |  |  |  | 1 | .20 | .24 | .26 | .16 | -.30 | -.33 |
| RCFT Copy |  |  |  |  |  |  |  |  |  |  | 1 | .48 | .47 | .24 | -.21 | -.31 |
| RCFT IR |  |  |  |  |  |  |  |  |  |  |  | 1 | .91 | .41 | -.25 | -.32 |
| RCFT DR |  |  |  |  |  |  |  |  |  |  |  |  | 1 | .38 | -.24 | -.33 |
| RCFT Rec |  |  |  |  |  |  |  |  |  |  |  |  |  | 1 | -.13 | -.22 |
| TMT A |  |  |  |  |  |  |  |  |  |  |  |  |  |  | 1 | .52 |
| TMT B |  |  |  |  |  |  |  |  |  |  |  |  |  |  |  | 1 |

*Note.* RAVLT = Rey Auditory Verbal Learning Test. RAVLT 1-5 = Total words across five learning trials. RAVLT A6 = immediate recall (after distractor list). RAVLT A7 = delayed recall (after 20 min. delay). RAVLT Rec = recognition discrimination (hits – false alarms). NSF = Number Span Forward. NSB = Number Span Backward. JOLO = Judgement of Line Orientation. F Flu = phonemic fluency for words starting with “F” (from the Montral Cognitive Assessment). L Flu = phonemic fluency for words starting with “L”. Cat Flu = category/semantic fluency for animals. RCFT = Rey Complex Figure Test. RCFT Copy = copy trial. RCFT IR = immediate recall trial. RCFT DR = delayed recall trial (after 30 min. delay). RCFT Rec = recognition discrimination (hits – false alarms). TMT A = Trail Making Test Part A. TMT B = Trail Making Test Part B.

Eigenvalues are presented in the scree plot in **Figure 1**. Based on Kaiser’s rule and examination of the scree plot, five factors should be retained. Parallel analysis was performed using 1000 simulated datasets, which also suggested retaining five factors. We found the five-factor model met our criteria of having every variable load onto a single factor with a loading greater than 0.30. The five-factor model explained 56% of the total variance. **Table 3** presents the factor loadings for the five-factor model. The factors can be characterized by the underlying cognitive domains: Factor 1 = Verbal Memory (RAVLT learning, immediate recall, delayed recall, recognition discrimination), Factor 2 = Visual Memory (RCFT copy, immediate recall, delayed recall, recognition discrimination), Factor 3 = Fluency (F and L letter fluency, animal category fluency), Factor 4 = Working Memory (NSF, NSB), and Factor 5 = Visual Processing (JOLO, TMT A & B). Several factors were moderately to strongly correlated (see **Table 4**), suggesting an oblique rotation was appropriate for these data.

**Table 3.** Factor Loadings.

| **Variable** | **F1** | **F2** | **F3** | **F4** | **F5** |
| --- | --- | --- | --- | --- | --- |
| **RAVLT A1-5** | 0.82 | -0.01 | 0.02 | 0.10 | 0.01 |
| **RAVLT A6** | 0.92 | 0.00 | -0.04 | 0.01 | 0.01 |
| **RAVLT A7** | 0.94 | 0.00 | 0.01 | -0.05 | -0.02 |
| **RAVLT Rec** | 0.69 | 0.03 | -0.01 | -0.05 | -0.01 |
| **RCFT Copy** | 0.02 | 0.43 | -0.02 | -0.01 | 0.21 |
| **RCFT IR** | 0.00 | 0.99 | 0.01 | -0.01 | -0.07 |
| **RCFT DR** | 0.00 | 0.96 | 0.02 | -0.01 | -0.05 |
| **RCFT Rec** | 0.03 | 0.38 | -0.03 | 0.01 | 0.08 |
| **F Fluency** | -0.02 | -0.02 | 0.80 | -0.03 | -0.06 |
| **L Fluency** | -0.03 | -0.01 | 0.76 | 0.05 | 0.02 |
| **Category Fluency** | 0.14 | 0.06 | 0.33 | -0.02 | 0.22 |
| **NSF** | -0.06 | -0.03 | 0.04 | 0.68 | -0.03 |
| **NSB** | 0.06 | 0.03 | -0.03 | 0.69 | 0.00 |
| **JOLO** | -0.07 | 0.22 | -0.05 | 0.11 | 0.36 |
| **TMT A** | -0.01 | 0.02 | 0.00 | 0.07 | -0.88 |
| **TMT B** | 0.02 | -0.04 | 0.00 | -0.01 | -0.76 |

*Note.* RAVLT = Rey Auditory Verbal Learning Test. RAVLT 1-5 = Total words across five learning trials. RAVLT A6 = immediate recall (after distractor list). RAVLT A7 = delayed recall (after 20 min. delay). RAVLT Rec = recognition discrimination (hits – false alarms). RCFT = Rey Complex Figure Test. RCFT Copy = copy trial. RCFT IR = immediate recall trial. RCFT DR = delayed recall trial (after 30 min. delay). RCFT Rec = recognition discrimination (hits – false alarms). NSF = Number Span Forward. NSB = Number Span Backward. JOLO = Judgement of Line Orientation. TMT A = Trail Making Test Part A. TMT B = Trail Making Test Part B. F1 = Factor 1 (Verbal Memory), F2 = Factor 2 (Visual Memory), F3 = Factor 3 (Fluency), F4 = Factor 4 (Working Memory), F5 = Factor 5 (Visual Processing).

**Table 4.**
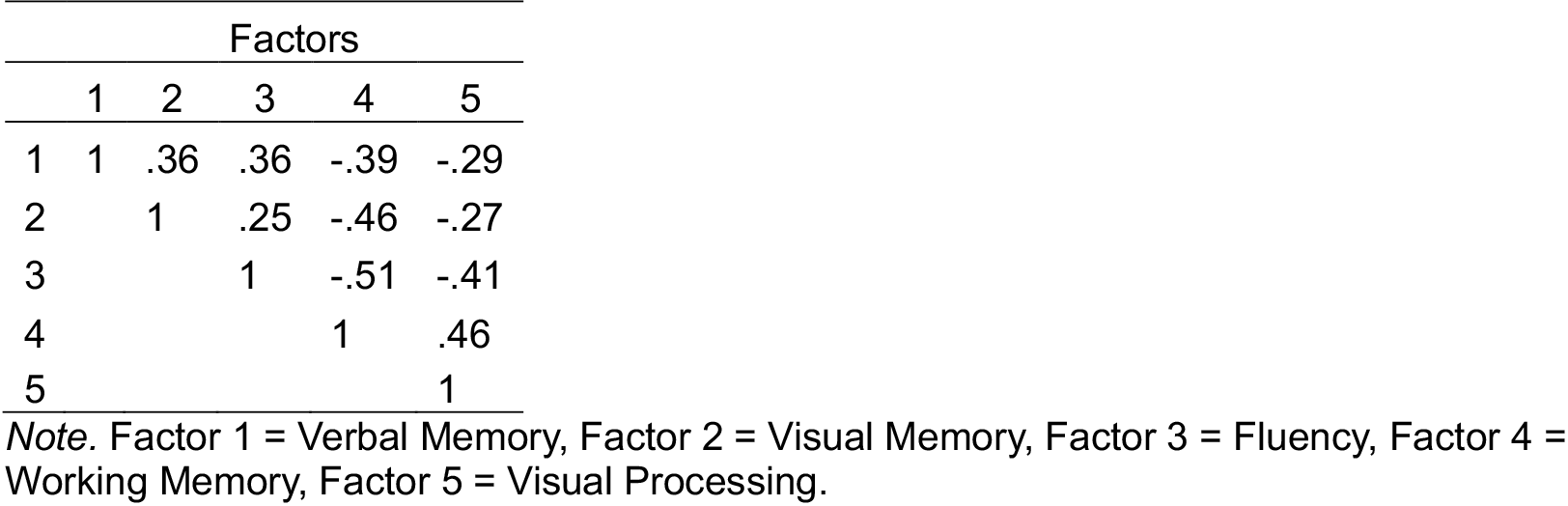
Factor Intercorrelations.

**Figure 1.**
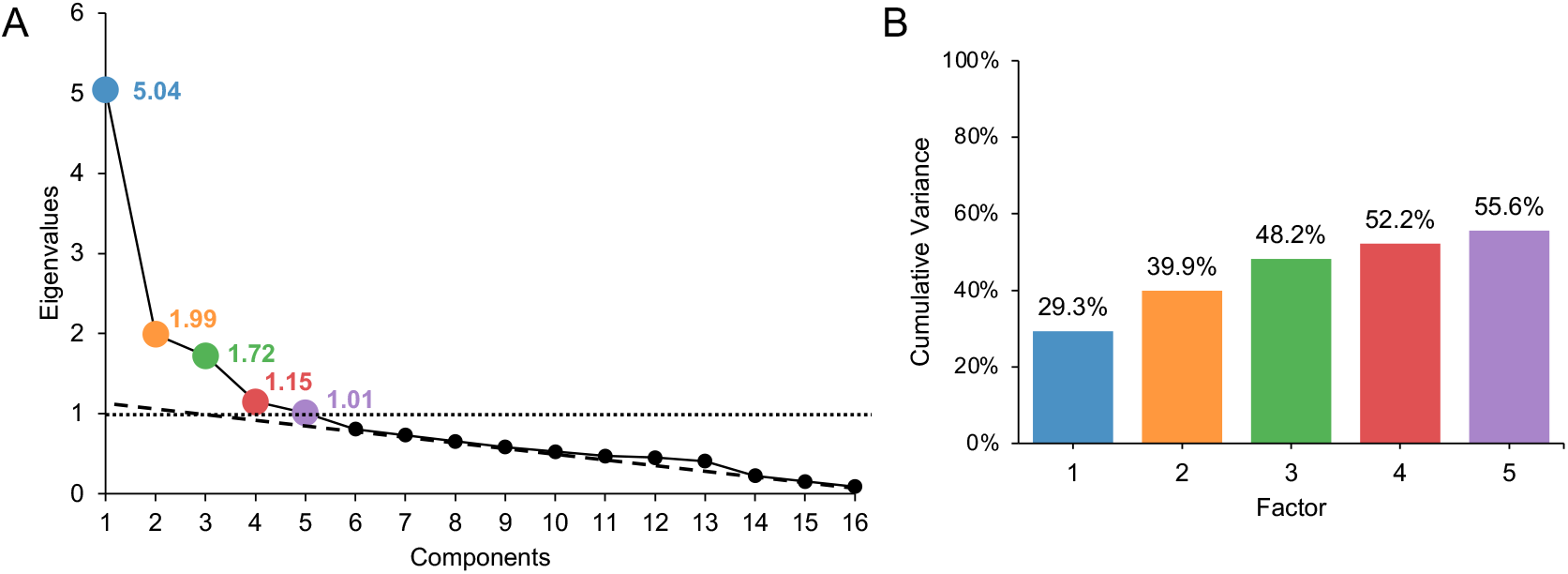
Scree Plot and Cumulative Variance Explained. In the scree plot (**A**), the dotted line depicts the cutoff for Kaisser’s rule (> 1) and the dashed line depicts the scree plot’s optimal coordinates index. Five eigenvalues meet the criteria for Kaiser’s rule, scree plot, and parallel analysis. (**B**) Bar graph showing cumulative variance explained by each of the five factors.

### 3.2. Relationship Between Factor Scores and Structural MRI, Functional Networks, and CSF Biomarkers

Of the 1,697 participants with factor scores and *APOE* data, 1,257 (74.1%) had CSF data, 1,208 (71.2%) had structural MRI data, and 825 (48.6%) had functional connectivity MRI data. All results are presented controlling for demographics and *APOE* genotype.

We found that Factor 1 and Factor 2 were positively associated with hippocampal volume (β = 0.09, 95% CI: [0.01, 0.16], *p* = .025 and β = 0.11, [0.04, 0.18], *p* = .002, respectively; **Figure 2A**). No other factors were associated with hippocampal volume (*p*s > .221), and no factors were associated with entorhinal cortex volume (*p*s > .051).

**Figure 2.**
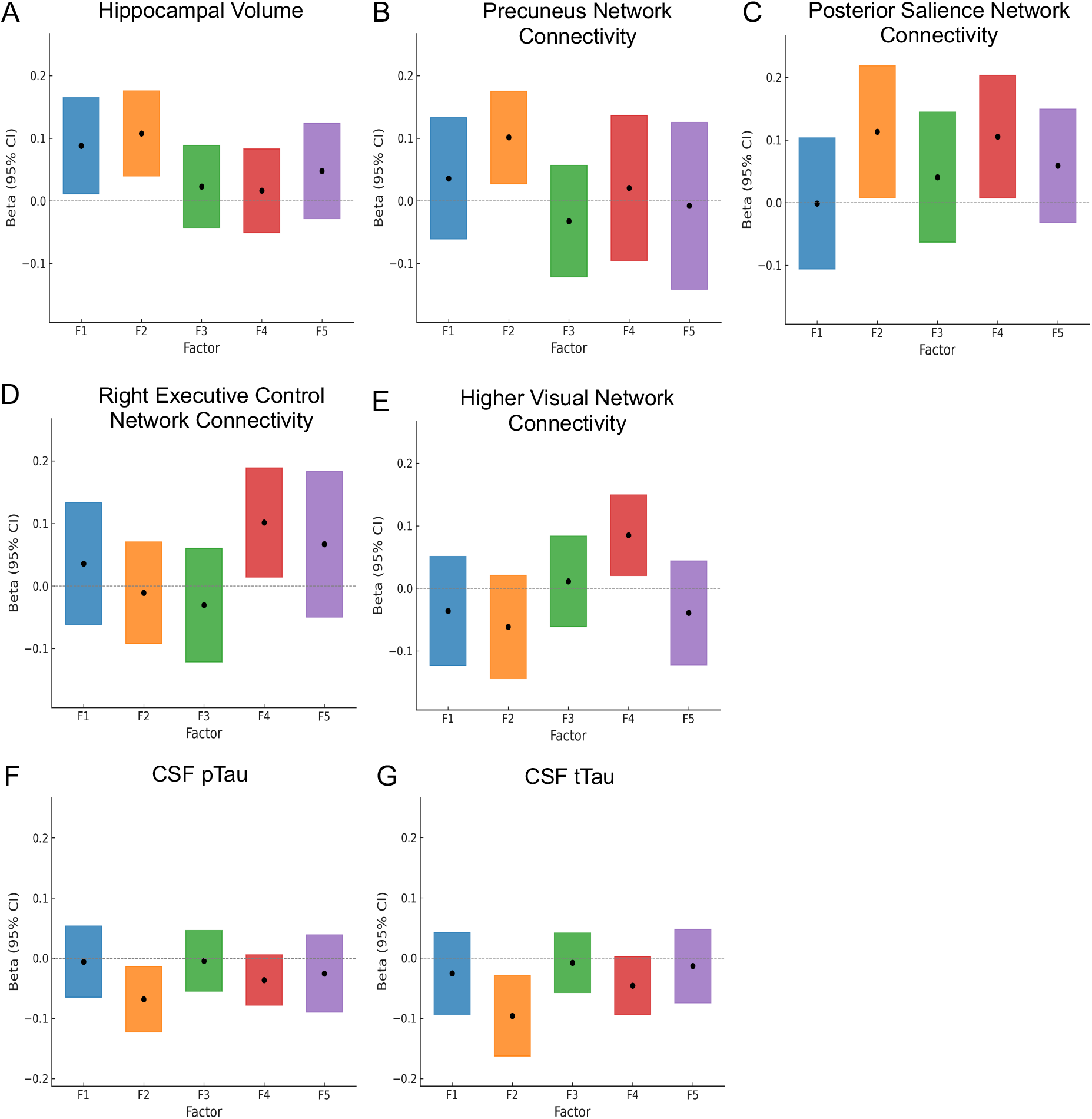
Associations Between Factor Scores and Hippocampal Volume (A), Functional Connectivity (B-E), and CSF pTau and tTau (F, G). Factor 1 = Verbal Memory, Factor 2 = Visual Memory, Factor 3 = Fluency, Factor 4 = Working Memory, Factor 5 = Visual Processing.

We found significant relationships between Factor 2 and connectivity in the precuneus (β = 0.10, [0.03, 0.18], *p* = .008) and posterior salience (β = 0.11, [0.01, 0.22], *p* = .036) networks; and between Factor 4 and connectivity in RECN (β = 0.10, [0.01, 0.19], *p* = .023), posterior salience (β = 0.11, [0.01, 0.20], *p* = .036) and higher visual network (β = 0.08, [0.02, 0.15], *p* = .010). No other relationships were significant (all *p*s > .056). These results are shown in **Figure 2B-E**.

Only Factor 2 was significantly associated with CSF levels of pTau (β = −0.07, [−0.86, - 0.08], *p* = .014; **Figure 2F**) and tTau (β = −0.10, [−11.9, −2.02], *p* = .005; **Figure 2G**; all other factors: *p*s > .089 for pTau, *p*s > .062 for tTau). No factors were associated with Aβ42 levels (*p*s > .091).

After multiple comparison adjustment with the Benjamini-Hochberg procedure to control the FDR, all p-values for the significant associations presented above were less than the corresponding Benjamini-Hochberg critical values and therefore considered statistically significant. Additionally, when ranking the p-values for the factor associations with tTau, the p-value for Factor 4 (β = −0.05, [−6.88, 0.25], *p* = .062) was less than the corresponding critical value (.08).

To determine which cognitive test scores from each factor made the strongest contributions in the significant relationships detailed above, we conducted follow-up median regression tests using the cognitive scores as predictor variables, demographics and *APOE* genotype as covariates, and the hippocampal volume, functional connectivity, and CSF data as outcome variables. The same FDR corrections were applied. Results are presented in **Table 5**. The relationship of Factor 2 to each significant outcome variable was driven primarily by RCFT immediate and delayed recall with less significant contributions from copy and recognition.

**Table 5.** Results of Follow-Up Tests.

| | Test | $\beta$ | 95% CI | $p$ | B-H Critical Value |
| --- | --- | --- | --- | --- | --- |
| Hippocampal Volume |  |  |  |  |  |
| Factor 1<br>(Verbal Memory) | RAVLT A1-5 | 0.061 | [-0.024, 0.145] | **.158** | .20 |
|  | RAVLT A6 | 0.075 | [0.003, 0.147] | **.043** | .15 |
|  | RAVLT A7 | 0.085 | [0.014, 0.155] | **.019** | .10 |
|  | RAVLT Rec | 0.118 | [0.030, 0.207] | **.009** | .05 |
| Factor 2<br>(Visual Memory) | RCFT Copy | -0.017 | [-0.096, 0.063] | .682 | .20 |
|  | RCFT IR | 0.107 | [0.036, 0.178] | **.003** | .10 |
|  | RCFT DR | 0.107 | [0.036, 0.179] | **.003** | .05 |
|  | RCFT Rec | 0.054 | [-0.003, 0.111] | **.066** | .15 |
| Precuneus Network Functional Connectivity |  |  |  |  |  |
| Factor 2<br>(Visual Memory) | RCFT Copy | 0.036 | [-0.037, 0.110] | .334 | .20 |
|  | RCFT IR | 0.096 | [0.018, 0.174] | **.016** | .05 |
|  | RCFT DR | 0.090 | [0.005, 0.175] | **.039** | .15 |
|  | RCFT Rec | 0.082 | [0.013, 0.151] | **.020** | .10 |
| Posterior Salience Network Functional Connectivity |  |  |  |  |  |
| Factor 2<br>(Visual Memory) | RCFT Copy | -0.002 | [-0.101, 0.097] | .971 | .20 |
|  | RCFT IR | 0.107 | [0.007, 0.208] | **.037** | .05 |
|  | RCFT DR | 0.045 | [-0.063, 0.153] | .412 | .15 |
|  | RCFT Rec | -0.039 | [-0.131, 0.052] | .400 | .10 |
| Factor 4<br>(Working Memory) | NSF | 0.110 | [0.027, 0.194] | **.010** | .10 |
|  | NSB | 0.017 | [-0.081, 0.115] | .735 | .20 |
| Right Executive Control Network Functional Connectivity |  |  |  |  |  |
| Factor 4<br>(Working Memory) | NSF | 0.093 | [0.015, 0.171] | **.020** | .10 |
|  | NSB | 0.077 | [-0.001, 0.156] | **.054** | .20 |
| Higher Visual Network Functional Connectivity |  |  |  |  |  |
| Factor 4<br>(Working Memory) | NSF | 0.031 | [-0.032, 0.094] | .339 | .20 |
|  | NSB | 0.099 | [0.027, 0.170] | **.007** | .10 |
| CSF pTau |  |  |  |  |  |
| Factor 2<br>(Visual Memory) | RCFT Copy | -0.001 | [-0.073, 0.070] | .968 | .20 |
|  | RCFT IR | -0.069 | [-0.126, -0.012] | **.018** | .10 |
|  | RCFT DR | -0.067 | [-0.120, -0.014] | **.014** | .05 |
|  | RCFT Rec | -0.026 | [-0.089, 0.037] | .418 | .15 |
| CSF tTau |  |  |  |  |  |
| Factor 2<br>(Visual Memory) | RCFT Copy | -0.015 | [-0.082, 0.051] | .652 | .15 |
|  | RCFT IR | -0.094 | [-0.155, -0.033] | **.003** | .10 |
|  | RCFT DR | -0.092 | [-0.150, -0.034] | **.002** | .05 |
|  | RCFT Rec | 0.0005 | [-0.064, 0.065] | .989 | .20 |

*Note.* p-values in bold text indicate those that are less than their respective Benjamini-Hochberg (B-H) critical values. RAVLT = Rey Auditory Verbal Learning Test. RAVLT 1-5 = Total words across five learning trials. RAVLT A6 = immediate recall (after distractor list). RAVLT A7 = delayed recall (after 20 min. delay). RAVLT Rec = recognition discrimination (hits – false alarms). RCFT = Rey Complex Figure Test. RCFT Copy = copy trial. RCFT IR = immediate recall trial. RCFT DR = delayed recall trial (after 30 min. delay). RCFT Rec = recognition discrimination (hits – false alarms). NSF = Number Span Forward. NSB = Number Span Backward.

## 4. Discussion

The objective of the current study was to determine whether performance on neuropsychological tests could predict early changes in AD biomarkers in cognitively normal individuals. Using exploratory factor analysis, we grouped neuropsychological test scores into distinct cognitive domains and examined their relationship with structural MRI, resting-state functional connectivity, and CSF Aβ42, tTau, and pTau. Our major findings involved Factor 2, which consisted of scores on the RCFT—a measure of visuospatial construction and visual episodic memory. Higher scores on this factor were significantly associated with larger hippocampal volume, greater functional connectivity in the precuneus and posterior salience networks, and lower CSF tTau and pTau levels. Follow-up tests showed that RCFT immediate and delayed recall scores were the main drivers of these relationships. These findings suggest that RCFT, particularly the recall elements, may be sensitive to certain biomarkers that have been associated with disease development years before cognitive decline is manifest.

### 4.1. Structural MRI

We found that both Factor 1 (Verbal Memory) and Factor 2 (Visual Memory) were positively associated with larger hippocampal volume. The hippocampus is one of the earliest sites of neurodegeneration in AD, with volumetric reductions often preceding clinical diagnosis by years.^13^ Our findings align with prior research showing that hippocampal atrophy is associated with verbal^46,47^ and visual^48^ memory impairments in individuals at risk for AD. Both the RAVLT and RCFT require encoding, storage, and retrieval of detailed information— processes that are critically dependent on hippocampal integrity. Identification of a significant relationship of cognition with a neurodegeneration marker (i.e., hippocampal volume) in a cognitively unimpaired cohort was unexpected and may be driven by presence of individuals experiencing the very earliest stages of cognitive decline.

### 4.2. Resting State Functional Connectivity

Our findings showed that Factor 2 (Visual Memory) was associated with greater resting state functional connectivity in the precuneus and posterior salience networks. The precuneus plays a central role in episodic memory retrieval and visuospatial imagery,^49^ and is among the earliest regions to accumulate amyloid and exhibit metabolic and connectivity disruptions in AD.^50,51^ Reduced precuneus network connectivity has been linked to CSF biomarkers of AD, even in asymptomatic individuals.^52^ Similarly, the posterior salience network, which supports attentional reorienting and integration of salient environmental stimuli,^53^ is also vulnerable in aging and AD progression.^54^ Our findings suggest that individuals with preserved visuospatial episodic memory exhibit stronger functional connectivity within these networks, potentially indicating resilience to early neurodegenerative changes.

We also observed significant associations between Factor 4 (Working Memory) and stronger connectivity in the right executive control (RECN), posterior salience, and higher visual networks. This finding is consistent with prior work^55^ showing that executive function relies on the integrity of the prefrontal cortex and its connectivity with large-scale networks. The RECN, which includes right dorsolateral prefrontal and parietal regions, is implicated in attentional control, task switching, and working memory. Stronger functional connectivity within these networks among individuals with better working memory performance may indicate intact or even upregulated network function, supporting the notion of network-based cognitive reserve.

### 4.3. CSF

An important aspect of our study was the relationship between cognitive performance and CSF biomarkers. We found that higher Factor 2 (Visual Memory) scores were associated with lower CSF tTau and pTau levels but not with Aβ42. Although Aβ42, tTau, and pTau are all considered early biomarkers of AD, prior studies suggest amyloid accumulation alone is insufficient to cause cognitive impairment.^56^ In contrast, abnormal tau accumulation appears to be more strongly associated with clinical progression in asymptomatic individuals at risk for AD.^57-59^ Similarly, our findings suggest that CSF tTau and pTau levels are more strongly associated with cognitive performance among unimpaired individuals than levels of Aβ42.

### 4.4. *APOE* Genotype

*APOE ε4* is the strongest genetic risk factor for late-onset AD and strongly influences levels of Aβ42, tTau, and pTau in cognitively normal individuals across the lifespan.^60^ While *APOE* genotype was a significant covariate for all CSF biomarkers, the presence or absence of an *ε4* allele was not associated with significant differences in any factor scores.

### 4.5. Conclusions and Future Directions

Overall, our results suggest that visual episodic memory, as evaluated by the RCFT, may be particularly sensitive to early AD biomarker changes within a cohort of cognitively unimpaired individuals. It is noteworthy that identified associations of Factor 2 (Visual Memory) with CSF tTau and pTau levels and precuneus and posterior salience network connectivity was not found with Factor 1 (Verbal Memory). While episodic memory is broadly recognized and incorporated as an important measure in AD-related studies and trials, this is most commonly assessed with verbal episodic memory tasks. Lack of use of the RCFT is likely due in part to the test being difficult and time-consuming to score. Loring et al.^61^ developed a 10-element recall scoring system and recognition task showing high correlation with the original scoring methodology while significantly reducing time burden. This simplified approach may lead to wider adoption of the RCFT which may be more effective at identifying individuals earlier in the evolution of AD pathology.

While our study provides valuable insights into the relationship of specific cognitive domains with biomarkers of AD, several limitations should be noted. First, our analyses were cross-sectional, limiting our ability to infer causality. As EHBS is a longitudinal study, our next step will be to incorporate data from subsequent visits to track cognitive and neurobiological changes over time. Second, the sample was of an unimpaired population, with limited variability in cognitive performance and MRI data. Third, AD biomarkers were limited to CSF and MRI features and amyloid and tau PET results were not included. Finally, our analysis lacks well-designed task-based fMRI, which may be more sensitive to early changes in AD. Outstanding questions include: 1) Do other visuospatial episodic memory tasks show similar sensitivity as RCFT to AD biomarkers in asymptomatic individuals? 2) Does the factor structure change in subsequent testing with the influence of practice effects? 3) How does the factor structure hold up in clinical populations? 4) Do other factors become more relevant further into disease progression? Future research should continue to explore the role of visuospatial episodic memory in identifying individuals at risk for cognitive decline, with an emphasis on longitudinal assessment in diverse populations.

## Data Availability

All data produced in the present study are available upon reasonable request to the authors

## 5. Acknowledgments

We are grateful for all Emory Healthy Brain Study participants who generously provided data and biospecimens for the study. We acknowledge the efforts of staff who support the Emory Goizueta ADRC, EHBS, and the Emory Cognitive Neurology Clinic, without whom this work would not be possible. Staff and investigators in the Emory Healthy Aging Study and Emory Healthy Brain Study can be found at: https://healthyaging.emory.edu/team and https://healthyaging.emory.edu/team/study-staff/.

